# Influence of body mass index on SAPS3 prognostic performance in critically ill patients from Brazil

**DOI:** 10.1101/2020.04.30.20086058

**Authors:** Isabella B. B. Ferreira, Rodrigo C. Menezes, Matheus L. Otero, Thomas A. Carmo, Gabriel P. Telles, Bruno V. B. Fahel, Manoel Barral-Netto, Maria B. Arriaga, Kiyoshi F. Fukutani, Licurgo Pamplona Neto, Gabriel A. Agareno, Sydney Agareno, Kevan M. Akrami, Nivaldo M.Filgueiras Filho, Bruno B. Andrade

## Abstract

Obesity has emerged as a significant global health problem, and its association with increased morbidity and mortality is well established. An obesity paradox has been extensively documented in the critically ill, appearing as a protective factor. Whether body mass index (BMI) impacts critical care severity scores has not been extensively studied previously, particularly in developing countries. This study aimed to evaluate the performance of severity scores across different BMI categories in a tertiary intensive care unit in Brazil. Observational and analytical cohort study in a general ICU in Northeastern Brazil between August 2015 and July 2018 that included all patients over 18 years of age admitted to the ICU. A total of 2,179 patients were included, with a mean age of 67.9 years and female predominance (53.1%). Similar to previous findings, those with overweight and obesity of any grade were not significantly associated with mortality, though for each additional 1kg/m2 there was a decrease of 0.04% in odds of death. The Simplified Acute Physiology Score III (SAPS3) accurately predicted mortality in all groups except in those underweight. Low weight appeared as an independent risk factor for mortality in the ICU. Furthermore, this is the first study to identify poor prognostic performance of a common ICU severity score in those with low weight, highlighting the need for alternative more precise metrics.

## Introduction

The number of obese individuals has tripled since 1975, affecting nearly 13% of the world’s adult population^1^. In Brazil, obesity affects one in five adults^2^.

With rising prevalence of obesity in the general population, there will invariably be a higher proportion of obese patients in intensive care units (ICU)^3^. Multiple studies previously noted an inverse relationship between obesity and mortality in those with critical illness^4,5^. Moreover, given the influence of obesity on mortality, it is unclear whether existing prognostic scores perform well across all ranges of body mass index. A recent retrospective study in the United States found that inclusion of body mass index (BMI) as a variable in ICU severity scores did not improve discriminative function^6^. As such, it is critical to determine how prognostic scores perform in a Brazilian ICU population. To determine performance of a commonly used ICU severity score across BMI categories, we evaluated a large cohort of hospitalized patients in a general ICU in Northeastern Brazil.

## Results

During the study period, 2,401 patients were admitted to the ICU, 222 individuals without data were excluded yielding a final cohort of 2,179 patients (Figure 1). Significant differences when stratified by BMI groups included older age, longer length of stay both in the ICU and prior to ICU admission in the underweight group. Clinically relevant differences at time of ICU admission included lower mean arterial pressure and increased C-reactive protein in the underweight group (Table 1).

**Figure 1.**
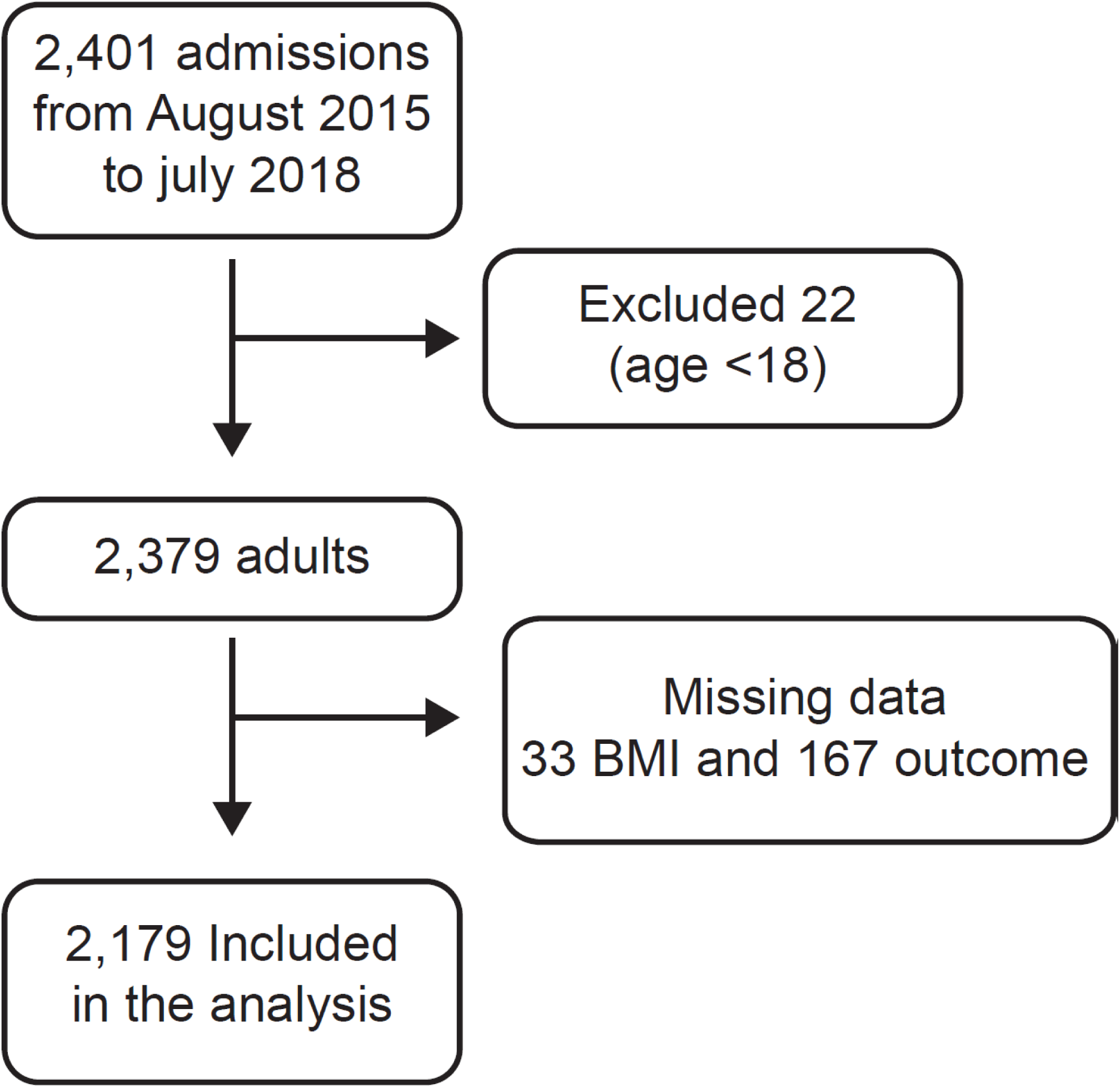
Flowchart of patients included in the study.

**Table 1.** Study population characteristics.

| Population Characteristics | Underweight<br>(n = 171) | Eutrophic<br>(n = 914) | Overweight<br>(n= 700) | Obese Grade I<br>(n = 272) | Obese Grade II/III<br>(n = 122) | p-value |
| --- | --- | --- | --- | --- | --- | --- |
| **Age (mean, SD)** | 76.06 ± 16.39 | 69.48 ± 18.51 | 66.10 ± 16.73 | 64.67 ± 16.19 | 62.40 ± 18.78 | < 0.0001 |
| **Gender, female (n, %)** | 98 (57.3) | 438 (47.9) | 356 (50.9) | 176 (64.7) | 90 (73.8) | < 0.0001 |
| **Admission Diagnosis (n, %).** |  |  |  |  |  | < 0.0001<sup>a</sup> |
| Cardiovascular | 25 (14.6) | 152 (16.6) | 160 (22.9) | 81 (29.8) | 34 (27.9) |  |
| Respiratory | 13 (7.6) | 55 (6.0) | 40 (5.7) | 13 (4.8) | 5 (4.1) |  |
| Neurological | 22 (12.9) | 171 (18.7) | 115 (16.4) | 41 (15.1) | 11 (9.0) |  |
| Infectious | 57 (33.3) | 184 (20.1) | 96 (13.7) | 30 (11.0) | 25 (20.5) |  |
| Surgical | 10 (5.8) | 141 (15.4) | 143 (20.4) | 65 (23.9) | 21 (17.2) |  |
| Other | 44 (25.7) | 211 (23.1) | 146 (20.9) | 42 (15.4) | 26 (21.3) |  |
| **Use of VAD** | 21 (12.3) | 86 (9.4) | 60 (8.6) | 27 (9.9) | 7 (5.7) | 0.375 |
| **Use of MV** | 32 (18.7) | 157 (17.2) | 110 (15.7) | 38 (14.0) | 10 (8.2) | 0.081 |
| **Length of stay prior to ICU (days)** | 4.95 ± 16.70 | 2.95 ± 14.58 | 1.65 ± 6.28 | 1.22 ± 4.77 | 1.08 ± 3.47 | 0.001 |
| **ICU length of stay (days)** | 9.77 ± 12.86 | 9.34 ± 15.85 | 6.84 ± 10.74 | 6.79 ± 12.07 | 5.58 ± 5.51 | < 0.0001 |
| **Congestive Heart Failure** | 11 (6.4) | 59 (6.5) | 44 (6.3) | 13 (4.8) | 5 (4.1) | 0.257<sup>a</sup> |
| **Chronic Renal Failure** | 17 (9.9) | 119 (13.0) | 82 (11.7) | 26 (9.6) | 7 (5.7) | 0.021<sup>a</sup> |
| **Cirrhosis** | 1 (0.6) | 9 (1.0) | 14 (2.0) | 3 (1.1) | 1 (0.8) | 0.453<sup>a</sup> |
| **Cancer** | 29 (17.0) | 149 (16.3) | 90 (12.9) | 23 (8.5) | 11 (9.0) | 0.004<sup>a</sup> |
| **Immunodeficient** | 2 (1.2) | 17 (1.9) | 4 (0.6) | 3 (1.1) | 1 (0.8) | 0.056<sup>a</sup> |
| **Diabetes Mellitus** | 61 (35.7) | 315 (34.5) | 277 (39.6) | 108 (39.7) | 56 (45.9) | 0.003<sup>a</sup> |
| **Coronary Artery Disease** | 12 (7.0) | 86 (9.4) | 106 (15.1) | 23 (8.5) | 14 (11.5) | 0.098<sup>a</sup> |
| **Stroke** | 41 (24.0) | 157 (17.2) | 96 (13.7) | 35 (12.9) | 11 (9.0) | 0.002 |
| **Dementia** | 29 (17.0) | 60 (6.6) | 25 (3.6) | 7 (2.6) | 4 (3.3) | <0.0001<sup>a</sup> |
| **Clinical and Laboratory (mean, SD)** |  |  |  |  |  |  |
| Mean arterial pressure (mmHg) | 91.88 ± 21.22 | 97.45 ± 20.80 | 99.73 ± 20.22 | 102.45 ± 20.71 | 101.32 ± 21.53 | < 0.0001 |
| Heart rate (bpm) | 89.70 ± 21.04 | 86.31 ± 20.83 | 84.46 ± 19.55 | 82.57 ± 19.17 | 87.90 ± 21.52 | 0.001 |
| Respiratory rate | 21.21 ± 4.98 | 20.10 ± 4.43 | 20.23 ± 4.34 | 19.83 ± 4.55 | 20.45 ± 4.21 | 0.021 |
| Creatinine(mg/dL) | 1.40 ± 2.27 | 1.49 ± 2.28 | 1.52 ± 2.63 | 1.26 ± 1.82 | 1.08 ± 1.10 | 0.249 |
| Platelets (x10<sup>3</sup>) | 252.5 ± 136.0 | 239.7 ± 113.7 | 227.3 ± 110.3 | 221.1 ± 76.0 | 259.8 ± 149.3 | 0.001 |
| Hematocrit | 32.34 ± 6.75 | 34.17 ± 7.58 | 35.20 ± 7.38 | 36.75 ± 6.65 | 36.10 ± 5.94 | < 0.0001 |
| Na (mEq/L) | 140.2 ± 8.87 | 138.7 ± 7.68 | 139.7 ± 6.25 | 140.0 ± 6.48 | 139.1 ± 5.40 | 0.014 |
| C Reactive Protein (mg/dL) | 96.77 ± 64.73 | 91.38 ± 70.15 | 79.53 ± 70.15 | 72.48 ± 67.35 | 73.09 ± 68.99 | < 0.0001 |
| **Mortality and Morbidity (n, %)** |  |  |  |  |  |  |
| **ICU deaths** | 64 (37.4) | 172 (18.8) | 76 (10.9) | 21 (7.7) | 10 (8.2) | < 0.0001 |

ICU = Intensive care unit; Simplified Acute Physiology Score III (SAPS3); Charlson Comorbidity Index (CCI);
VAD = Vasoactive drug; MV =Mechanical ventilation.
<sup>a</sup>Chi-square test.

A total of 343 (15.7%) deaths occurred in the ICU during the study period, a mortality rate comparable to that predicted by the SAPS3 score (13.3%). When mortality was stratified by weight, underweight participants compared to eutrophic had a higher rate of death than predicted by SAPS3 (37% vs 26.6%), higher admission SAPS3 (54.0 vs 48.5, p<0.0001) and MFI (1.94 vs 1.62 p=0.019) (Supplemental Table 1). In contrast, overweight, obese and advanced obese individuals had lower rates of death than expected by SAPS3 (10.9%, 7.7%, 8.2% vs 14.5%, 13.3%, 14.8%, respectively). The ability of SAPS3 to predict mortality was evaluated for each BMI group. We found that discriminate function was excellent in all groups except those underweight, with significantly decreased AUC reflecting decreased sensitivity and specificity for SAPS 3 performance (Figure 2).

**Figure 2.**
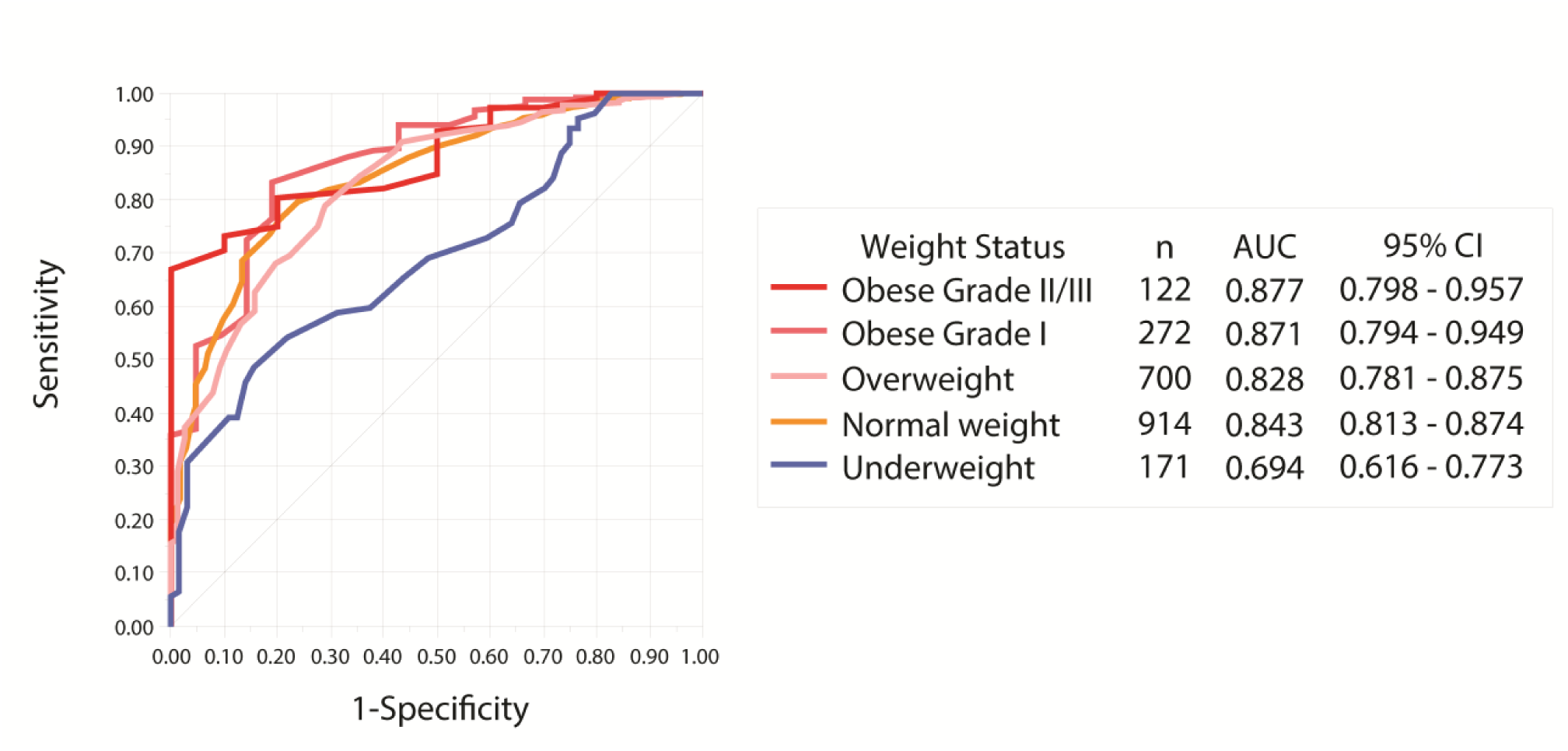
Performance of SAPS3 Mortality Prediction According to BMI Category. SAPS3 performance was accurate in all BMI groups except those in underweight, where significantly poor discriminate function was noted. P-value for all curves <0.001.

Univariate analysis demonstrated a significant increased odd of death 3.71 (95% CI, 2.65-5.18)] for underweight patients (Supplemental Figure 1). Overweight, obese and advanced obese patients demonstrated a significantly reduced odd of death [95% CI, 0.55 (042-0.70), 0.36 (0.22-0. 58) and 0.38 (0.19-0.75), p-value <0.0001, <0.0001 and 0.0038 respectively]. Cancer and stroke were comorbidities associated with mortality independent of BMI subgroup (Supplemental Table 2). The following factors identified in univariate analysis (Supplemental Table 2) were evaluated in a multivariate regression model to determine the impact on mortality: heart rate, respiratory rate, vasopressor use, length of stay prior to ICU, urea, creatinine, cancer, stroke, C-reactive protein use of mechanical ventilation and BMI category. Increased mortality persisted in the underweight group with an OR 3.50 (95% CI, 1.43-8.58, p=0.006), while the overweight and obese groups did not result in lower probability of death when adjusted for other variables in the multivariate model. Use of mechanical ventilation OR 3.11 (95% CI, 4.90-8.24, p<0.0001) and use of vasopressors OR 2.69 (95% CI, 1.74-4.18, p<0.0001) were variables that fit best with mortality in our model (Supplemental Figure 1). In a Cox regression model, the variables that result in survival differences amongst the study groups were: underweight, vasopressor use, cancer (Figure 3).

**Figure 3.**
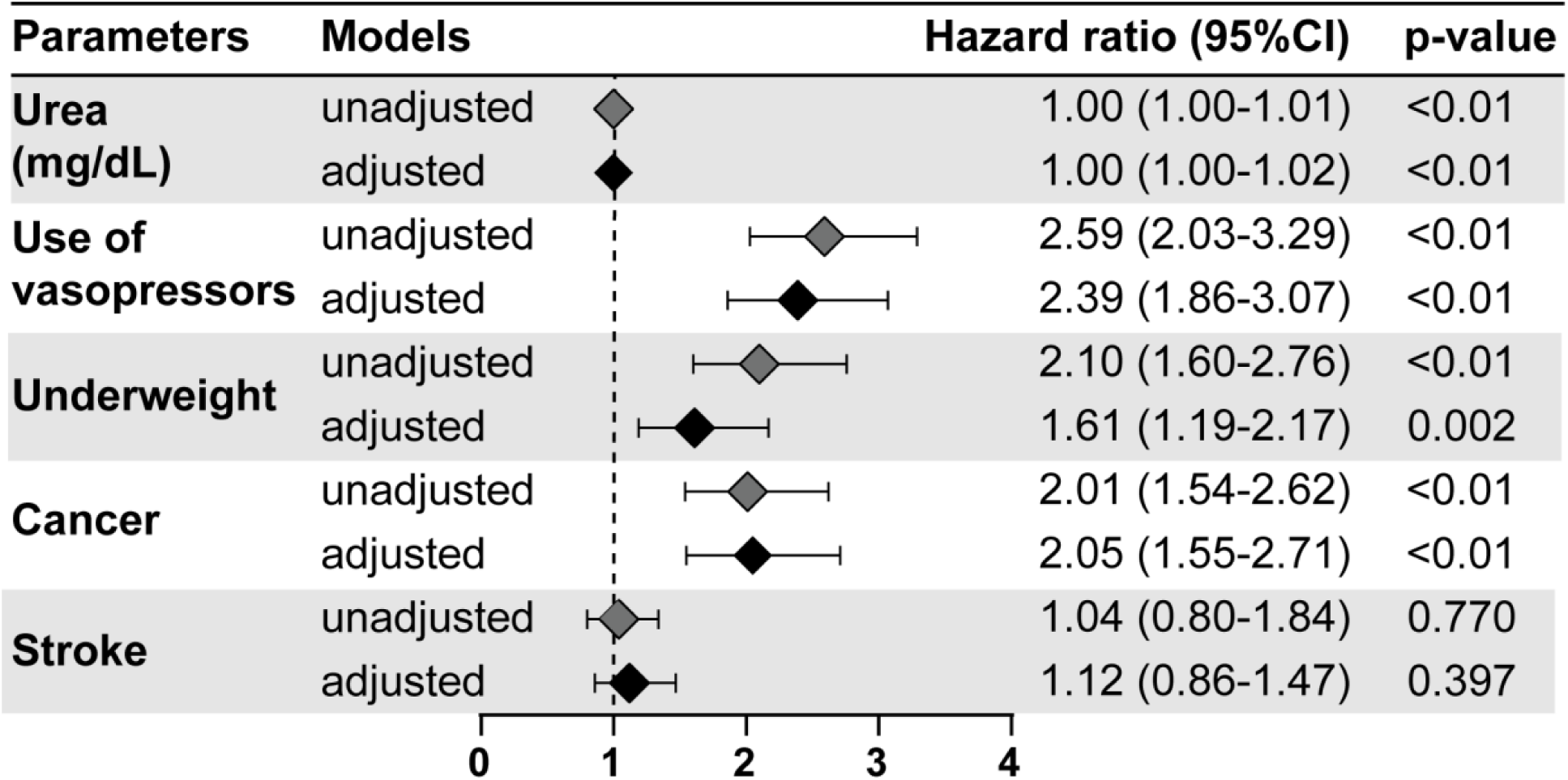
Cox Regression Analysis for ICU Mortality. Multivariate model yielded adjusted and unadjusted Hazard Ratios for variable with significance in univariate analysis.

A log-rank test was performed to determine whether differences in the survival distribution existed for the different types of study groups (underweight, overweight, obese I and obese II/III), using normal weight as the reference. After a median follow up of 32 days, the only statistically significant survival distribution was the comparison of underweight with normal weight, HR = 2.05 (95% 1.55-2.71), p <0.01. When the groups were stratified for SAPS3 score, only the obese II/III category lacked a significant difference when compared to those with high and low SAPS3 (considering a high score value > 50). An ICU survival analysis over the course of the study using a BMI cutoff of 18.5kg/m2 determined by ROC analysis demonstrated decreased survival in the underweight compared to the non-underweight population (Figure 4). Moreover, when evaluated as a continuous variable, the BMI presented statistical significance on both the univariate and multivariate analysis (p<0,0001 and p=0,011, respectively), with a decrease of 0.04% in the odds of mortality for each additional 1kg/m^2^ (Figure 5).

**Figure 4.**
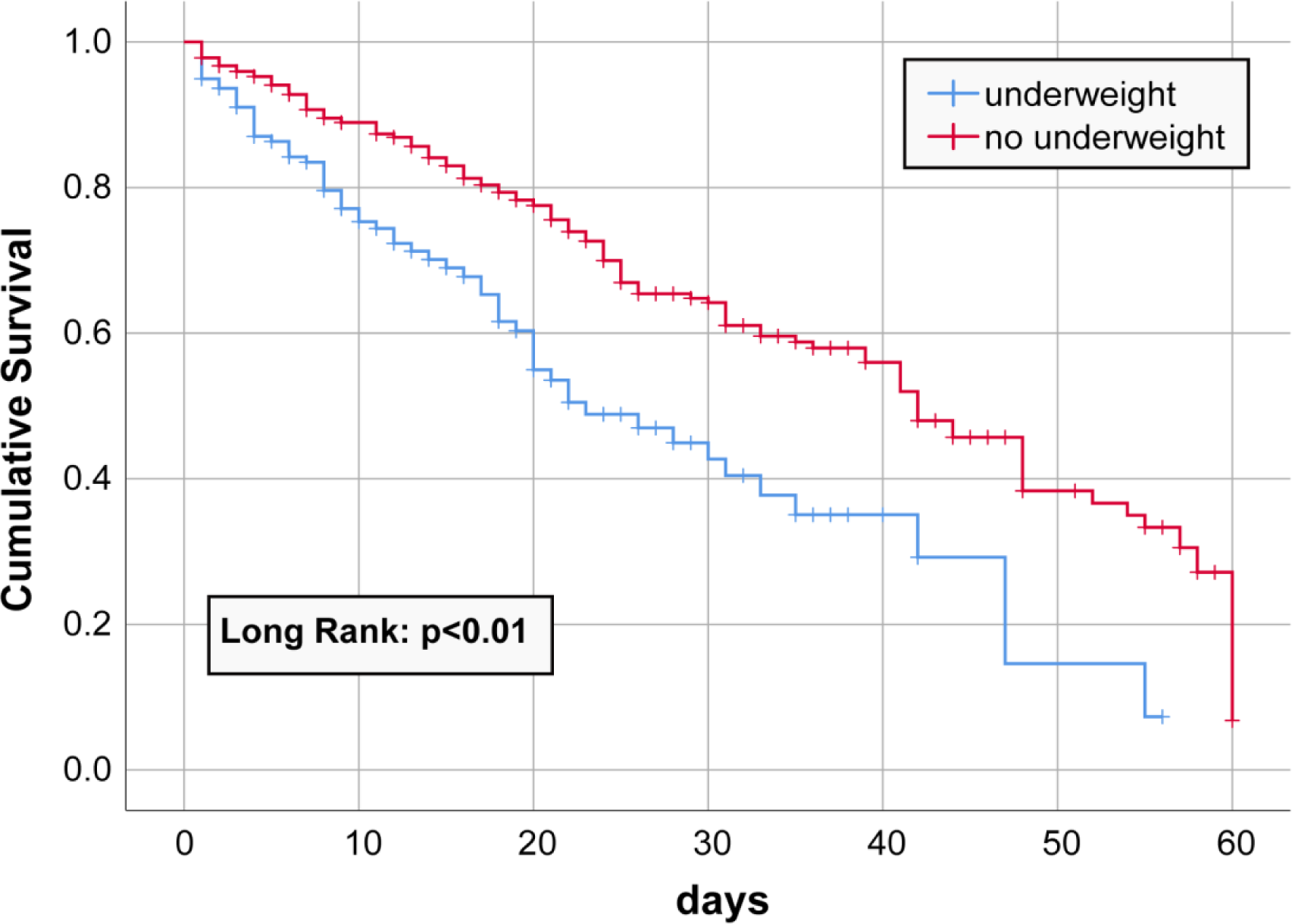
Kaplan Meier Survival Curve. A ROC curve analysis identified a BMI of 18.5kg/m^2^ as a cutoff that was used to stratify the cohort and evaluate survival over the study period.

**Figure 5.**
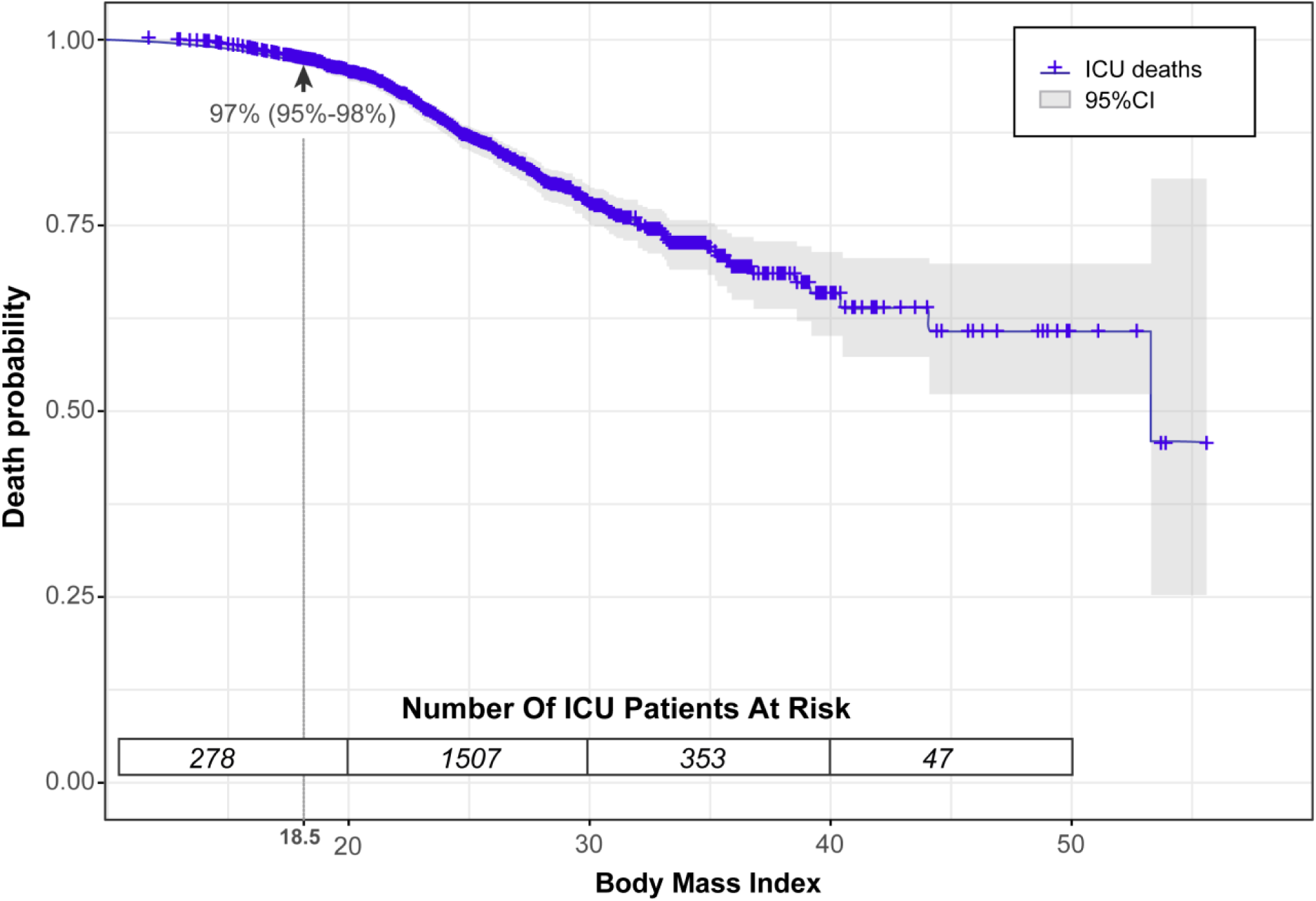
Death Probability According to BMI. For every 1kg/m^2^ there is a reduction of 0.04% probability of death.

## Discussion

This study is one of the first to evaluate, in Brazilian population, whether BMI affects accuracy of prognostic scores and outcomes in ICU.

Prognostic scores in critically ill individuals are intended to identify those most at risk of death during screening, particularly in resource-limited settings. In our study, while SAPS3 performed well in the overweight, obese, and eutrophic groups, there was a significant lack of prognostic discrimination in underweight patients. Similarly, Deliberato et al, 2019 demonstrated that the performance of the APACHE IV and OASIS severity scores were inconsistent across BMI categories^6^. In a prior study including only obese and eutrophic patients, it appeared that misclassification of prognostic scores, within the same severity of illness category, may result from primarily from differences in baseline laboratory markers in these two populations. It may be that this is may explain why severity scores performed poorly in underweight patients^6^.

Previous international research has demonstrated conflicting results regarding the association between BMI and mortality, with 4 observational studies finding that a BMI between 25-35kg/m^2^ was associated with greater short and medium term mortality^7–10^, while 14 studies and a metanalysis found no mortality difference based on BMI^11–23^ in contrast to 14 studies and two metanalysis finding increases in BMI to be protective in the ICU^3,16,24–35^. We determined that underweight individuals are at an increased risk of death and obesity may not be protective when adjusted for significant comorbidities including stroke and cancer.

Several hypotheses may explain why underweight, as opposed to overweight and obese, is a risk factor in this population, though there is a paucity of rigorous clinical evidence. Obese patients tend to be younger at the time of ICU admission, a population generally at lower mortality risk^5^. This was evident in our findings with a lower mean age among those with higher BMI. Alternatively, medical staff, anticipating possible risks and complications, may admit obese patients earlier to the ICU in relatively stable condition to initiate aggressive interventions^4^. In support of this hypothesis, obese individuals in our study were admitted with lower SAPS3 and CCI while underweight patients presented higher mean SAPS3 and MFI scores (Supplementary Table 1), albeit with poor predictive performance. Others have suggested that obese individuals have a greater nutritional reserve, thereby offering protection against hypercatabolic states experienced during critical illness as compared to their underweight counterparts^4,23^. Still others hypothesize that the adipokine profile in obese patients may modulate and dampen immunological response to severe acute illness which may be absent in the underweight population^4^. While we did not confirm obesity as a protective factor when adjusting for comorbidities, a mortality risk for underweight critically ill patients found in our study mirrors prior work in the developed world^23,24,32,34^. While there may be unknown host or environmental factors that differ between critically ill patients in Brazil and other developed countries, it is unlikely that this has influenced the findings that underweight status independently increases risk of death in the ICU.

Our study has certain limitations. First, as a single center there may be unmeasured residual confounders (ventilator settings, vasopressor doses, e.g.) that impacted mortality independent of BMI. The use of BMI as a parameter for obesity diagnosis, while useful at the population level, may not distinguish between lean mass and fat mass, thus being less precise in elderly and muscular individuals^36^. Moreover, BMI may not accurately assess visceral fat, a risk factor for disease independent of total body fat^17^. Future studies using BMI in conjunction with other methods to measure body fat may address this limitation. Height and weight measurements may have varied amongst healthcare professionals, compounded by possible differences in mean time elapsed between the weight assessment and ICU admission.

In conclusion, a greater chance of death was identified for underweight patients hospitalized in a general Brazilian ICU. Prognostic scores may overestimate risk of death in the obese while lacking discriminate function in predicting mortality in the underweight. Further studies will be needed to investigate whether this phenomenon persists in specific critically ill groups.

## Methods

### Ethics approval and consent to participate

All clinical investigations were conducted according to the principles expressed in the Declaration of Helsinki. Ethics approval and waiver of consent to participate was approved by the Research Ethics Committee of Hospital Ana Nery under the number 2.571.265 and CAAE 52892315.1.0000.0045.

### Clinical study design

Observational analytical cohort study conducted from August 2015 to July 2018 in a hospital with a general ICU of 22 beds in Salvador, Bahia, Brazil. All patients consecutively admitted to the ICU with length of stay longer than 24 hours were included. Patients under 18 years of age or those with missing data were excluded. Data was prospectively recorded in the Epimed Monitor system.

Covariates included: age, weight, height, sex, comorbidities, functional capacity, admission diagnosis, length of ICU and hospital stay, physiological and laboratorial data within the first 6 hours of admission, complications, use of supportive therapy in the ICU, ICU mortality and prognostic scores. We sought to determine whether mortality varied when stratified by body mass index (BMI) according to international standards recommended by WHO in underweight (BMI lower than 18.5kg/m^2^), normal weight (BMI between 18.5 and 24.99 kg/m^2^), overweight (BMI between 25 and 29.99 kg/m^2^), obese grade I (BMI between 30 and 34.99 kg/m^2^), obese grade II/III (BMI more than 35 kg/m^2^)^1^. We then evaluated the influence of BMI on the prognosis of critically ill patients, and examined whether or not the Simplified Acute Physiology Score (SAPS3) was able to predict ICU mortality accurately regardless of the BMI. Additionally, BMI was analyzed as a continuous variable to assess the analytical capacity of the variable. Through this analysis, we identified a BMI cutoff according to the receiver operator characteristics (ROC) curve analysis finding that a low weight BMI of 18.49 demonstrated the highest sensitivity for ICU mortality. This grouping was subsequently used in a Kaplan Meier survival analysis. The following Prognostic ICU scores were determined for all study patients: Modified Frailty Index (MFI), the SAPS3, the Charlson Comorbidity Index (CCI) without assigning points for age and Performance status, using a scale of zero to 2 based on the patient’s ability to perform daily living activities^37–40^.

### Data Analysis

Categorical variables were expressed as frequency and percentages, and continuous variables were expressed as mean (standard deviation, SD) for continuous variables with normal distribution, or median (interquartile range, IQR) for non-normal distribution, with normality being determined by Kolmogorov-Smirnov or Shapiro-Wilk test.

The proportion of categorical variables between groups were compared using the Fisher’s exact test or chi-squared. The mean/median of continuous variables were compared using independent T-tests, for parametric variables, or Mann-Whitney U test, for non-parametric variables, when analyzing the outcome groups, and also using one-way analysis of variance (ANOVA), for parametric variables, or Kruskal-Wallis, for non-parametric variables, when analyzing BMI categories. All tests were two-tailed and considered statistically significant for p< 0.05.

In order to assess for potential confounders, variables that demonstrated possible statistical associations (p < 0.1) with ICU mortality and BMI categories in univariate analysis were analyzed in multivariate models with calculation of odds ratio (OR) with a confidence interval (CI) of 95%. Binary logistic regression was used to identify characteristics independently associated with death in the ICU. Predictive performance of prognostic scores amongst the 5 groups was tested by calculating area under ROC curve with area under curve AUC>0.8 considered the best predictive test. With a Kaplan-Meier analysis, survival from 1 to 378 days was calculated according to the BMI stratification cutoff identified by the ROC analysis. Continuous variables were transformed into categorical variables (based on means) and, utilizing the Log-rank test, the survival curves were compared and considered statistically significant for p<0.05. We then applied the criteria of Hosmer Lemeshow: variables with p<0.1 in the Log-Rank test were eligible for the multivariate Cox model. To identify variables associated with survival, Hazard Ratios (HR) with confidence intervals were calculated. The data were analyzed with Microsoft Excel suite Office 365, GraphPad Prism version 6.01 and Statistical Package for the Social Sciences (SPSS), version 25.0.

## Data Availability

All the data used for analyses will be available after publication of the material upon request addressed to the corresponding author.

## Acknowledgments

Thank those who contributed directly or indirectly to the construction of this article, research groups GEMINI, linked to the Núcleo de Ensino e Pesquisa do Hospital da Cidade, and MONSTER, linked to the Osvaldo Cruz Foundation. The work of B.B.A. was supported by a grant from NIH (Rodrigo). I.B.B.F. was supported by UNEB: PICIN/UNEB. K.F.F. received a fellowship from the Programa Nacional de Pós-Doutorado, CAPES. The funders had no role in study design, data collection and analysis, decision to publish, or preparation of the manuscript. MBA was supported by a PhD fellowship from the Fundação de Amparo à Pesquisa do Estado da Bahia (FAPESB).

## Author’s Contributions

IBBF, KFF, MBA, BBA, KMA analyzed and interpreted the patient data regarding the impact of obesity in the ICU. RCM, GAA, GPT, MLO, TAC, BVBF performed chart review, data collection and manuscript review. MBN, LPN, SA provided study oversight and manuscript revision. NMFF, KMA and BBA were the major contributors to design, implementation and production of the manuscript. All authors read and approved the final manuscript.

## Data availability statement

The datasets used and/or analyzed during the current study are available from the corresponding author on reasonable request.

## Supplemental Material

**Content**

Table S1

Table S2

Figure S1

**Table S 1.** Distribution of prognosis scores and mortality by BMI Categories

| Variable | Underweight<br>(n = 171) | Eutrophic<br>(n = 914) | Overweight<br>(n= 700) | Obese<br>(n = 272) | Advanced<br>obese<br>(n = 122) | p-value |
| --- | --- | --- | --- | --- | --- | --- |
| ICU deaths | 64 (37.4) | 172 (18.8) | 76 (10.9) | 21 (7.7) | 10 (8.2) | < 0.0001 |
| CCI | 2.06 ± 1.90 | 1.75 ± 1.84 | 1.55 ± 1.68 | 1.28 ± 1.63 | 1.24 ± 1.50 | < 0.0001 |
| MFI | 1.94 ± 1.30 | 1.62 ± 1.30 | 1.68 ± 1.21 | 1.67 ± 1.06 | 1.75 ± 1.08 | 0.040 |
| SAPS3 | 54.06 ± 11.49 | 48.52 ± 12.68 | 44.63 ± 11.49 | 43.01 ± 11.87 | 44.02 ± 12.39 | < 0.0001 |
| BRADEN | 13.00 ± 3.48 | 14.61 ± 3.76 | 15.26 ± 3.66 | 14.93 ± 3.70 | 15.34 ± 3.62 | < 0.0001 |
| Performance status |  |  |  |  |  | < 0.0001 |
| Completely independent | 113 (66.1) | 778 (85.1) | 641 (91.6) | 245 (90.1) | 109 (89.3) |  |
| Partially independent | 18 (10.5) | 65 (7.1) | 39 (5.6) | 18 (6.6) | 8 (6.6) |  |
| Fully dependent | 40 (23.4) | 71 (7.8) | 20 (2.9) | 9 (3.3) | 5 (4.1) |  |
| Use of VAD | 21 (12.3) | 86 (9.4) | 60 (8.6) | 27 (9.9) | 7 (5.7) | 0.375 |
| Use of MV | 32 (18.7) | 157 (17.2) | 110 (15.7) | 38 (14.0) | 10 (8.2) | 0.081 |
| C reactive Protein (mg/L) | 96.77 ± 64.73 | 91.38 ± 70.15 | 79.53 ± 70.15 | 72.48 ± 67.35 | 73.09 ± 68.99 | < 0.0001 |
| Length of stay prior to ICU admission (days) | 4.95 ± 16.70 | 2.95 ± 14.58 | 1.65 ± 6.28 | 1.22 ± 4.77 | 1.08 ± 3.47 | 0.001 |
| ICU length of stay (days) | 9.77 ± 12.86 | 9.34 ± 15.85 | 6.84 ± 10.74 | 6.79 ± 12.07 | 5.58 ± 5.51 | < 0.0001 |
| ICU readmission | 20 (11.7) | 105 (11.5) | 51 (7.3) | 14 (5.1) | 10 (8.2) | 0.004 |

Results shown by number (%), mean ± standard deviation or median [25-75]. ICU = Intensive care unit; Modified Frailty Index (MFI); Simplified Acute Physiology Score III (SAPS3); Charlson Comorbidity Index (CCI); VAD = Vasoactive drug; MV =Mechanical ventilation

**Table S 2.** Univariate analysis of clinical and laboratory exams in the first 6 hours of ICU admission.

| Variable | Total<br>(n = 2179) | Deaths<br>(n = 343) | Non-Deaths<br>(n = 1836) | p-value |
| --- | --- | --- | --- | --- |
| Age (years) | 68 ± 18 | 77 ± 15 | 66 ± 18 | < 0.0001 |
| Gender, female | 1158 (53.1) | 172 (50.1) | 986 (53.7) | 0.239 |
| BMI | 25.6 ± 5.7 | 23.1 ± 5.5 | 26.0 ± 5.6 | < 0.0001 |
| Eutrophic | 914 (41.9) | 172 (50.1) | 742 (40.4) | 0.001 |
| Underweight | 171 (7.8) | 64 (18.7) | 107 (5.8) | < 0.0001 |
| Overweight | 700 (32.1) | 76 (22.2) | 624 (34.0) | < 0.0001 |
| Obese Grade I | 272 (12.5) | 21 (6.1) | 251 (13.7) | < 0.0001 |
| Obese Grade II/III | 122 (5.6) | 10 (2.9) | 112 (6.1) | 0.015 |
| CHF | 132 (6.1) | 16 (4.7) | 116 (6.3) | 0.268 |
| CRF | 251 (11.5) | 45 (13.1) | 206 (11.2) | 0.311 |
| Cirrhosis | 28 (1.3) | 6 (1.7) | 22 (1.2) | 0.430 |
| Cancer | 302 (13.9) | 71 (20.7) | 231 (12.6) | < 0.0001 |
| Immune deficiency | 27 (1.2) | 7 (2.0) | 20 (1.1) | 0.177 |
| Diabetes | 817 (37.5) | 132 (38.5) | 685 (37.3) | 0.716 |
| Coronary Artery Disease | 241 (11.1) | 31 (9.0) | 210 (11.4) | 0.223 |
| Chronic Atrial Fibrillation | 137 (6.3) | 32 (9.3) | 105 (5.7) | 0.015 |
| Stroke | 340 (15.6) | 77 (22.4) | 263 (14.3) | < 0.0001 |
| Dementia | 125 (5.7) | 48 (14.0) | 77 (4.2) | < 0.0001 |
| Tobacco | 164 (7.5) | 19 (5.5) | 145 (7.9) | 0.147 |
| Alcoholism | 119 (5.5) | 22 (6.4) | 97 (5.3) | 0.436 |
| Psychiatric disease | 169 (7.8) | 23 (6.7) | 146 (8.0) | 0.509 |
| Dyslipidemias | 201 (9.2) | 24 (7.0) | 177 (9.6) | 0.128 |
| Systolic arterial pressure<br>(mmHg) | 138 ± 29 | 130 ± 30 | 139 ± 28 | 0.846 |
| Diastolic Blood Pressure | 79 ± 20 | 75 ± 21 | 80 ± 20 | < 0.0001 |
| Mean Arterial Pressure | 99 ± 21 | 93 ± 22 | 100 ± 20 | < 0.0001 |
| HR (bpm) | 86 ± 20 | 94 ± 22 | 84 ± 20 | < 0.0001 |
| RR (bpm) | 20 ± 4 | 21 ± 5 | 20 ± 4 | < 0.0001 |
| Temperature (C) | 36 ± 1 | 36 ± 1 | 36 ± 1 | 0.692 |
| Urea mmol/L | 57 ± 47 | 86 ± 64 | 51 ± 42 | < 0.0001 |
| Creatinine mg/dL | 1.44 ± 2 | 1.86 ± 2 | 1.36 ± 2 | < 0.0001 |
| Platelets (x10<sup>3</sup>) | 235 ± 113 | 230 ± 130 | 236 ± 109 | 0.420 |
| Hematocrit | 34 ± 7 | 32 ± 8 | 35 ± 7 | < 0.0001 |
| Leukocytes total (x10<sup>3</sup>) | 11 ± 11 | 13 ± 7 | 11 ± 11 | 0.011 |
| Bands | 176.1 ± 511.6 | 304.6 ± 666.5 | 152.1 ± 473.5 | < 0.0001 |
| Segmented (x10<sup>3</sup>) | 9.26 ± 5.74 | 11.05 ± 6.40 | 8.92 ± 5.54 | < 0.0001 |
| Eosinophils | 123.5 ± 246.5 | 112.6 ± 286.0 | 125.5 ± 238.4 | 0.404 |
| Basophils | 18.6 ± 48.7 | 8.58 ± 27.55 | 20.47 ± 51.55 | < 0.0001 |
| Lymphocytes (x10<sup>3</sup>) | 1.69 ± 8.84 | 1.27 ± 1.04 | 1.77 ± 9.61 | 0.360 |
| Atypical | 9.78 ± 172.8 | 28.58 ± 390.26 | 6.26 ± 83.38 | 0.034 |
| Monocytes | 562.6 ± 390.2 | 521.38 ± 410.73 | 570.34 ± 385.85 | 0.040 |
| Na (mEq/L) | 139.3 ± 7.11 | 138.86 ± 9.56 | 139.4 ± 6.53 | 0.211 |
| K (mEq/L) | 4.32 ± 0.85 | 4.56 ± 1.07 | 4.28 ± 0.79 | < 0.0001 |
| Use of VAD | 201 (9.2) | 93 (27.1) | 108 (5.9) | < 0.0001 |
| Use of MV | 347 (15.9) | 149 (43.4) | 198 (10.8) | < 0.0001 |
| C-reactive Protein (mg/L) | 84.64 ± 69.73 | 136.75 ± 63.45 | 74.37 ± 66.25 | < 0.0001 |
| Length of stay prior to ICU<br>admission (days) | 2.37 ± 11.31 | 4.12 ± 11.20 | 2.05 ± 11.30 | 0.002 |
| ICU Duration (days) | 8.04 ± 13.30 | 16.08 ± 24.20 | 6.54 ± 9.31 | < 0.0001 |
| ICU readmission | 200 (9.2) | 49 (2.2) | 151 (8.2) | < 0.0001 |

Results expressed by number (%), mean ± standard deviation or median [25-75].
MAP= Mean arterial pressure; HR = heart rate; RR = Respiratory rate.

**Figure S1.**
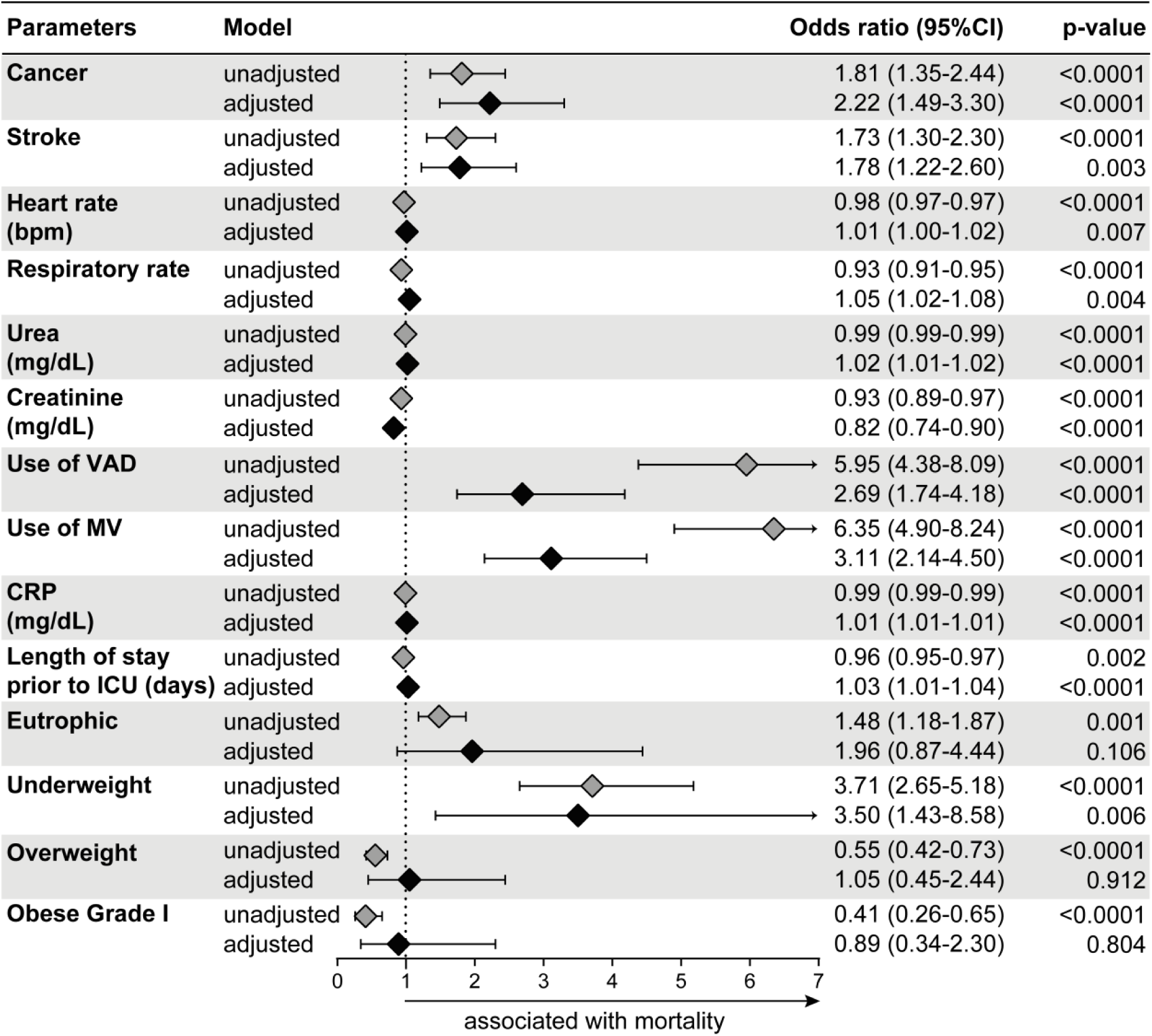
Adjusted and Unadjusted Multivariate Regression Model for ICU Mortality. Univariate analysis yielded unadjusted odds of death. Multivariate regression adjusted for differences in baseline characteristics (variables of p<0.1 identified in univariate analysis, Table S2).

